# Atovaquone for Treatment of COVID-19: A Prospective Randomized, Double-Blind, Placebo-Controlled Clinical Trial

**DOI:** 10.1101/2022.05.24.22275411

**Authors:** Mamta K. Jain, James A. de Lemos, Darren K. McGuire, Colby Ayers, Jennifer L. Eiston, Claudia L. Sanchez, Dena Kamel, Jessica A. Meisner, Emilia V. Thomas, Anita A. Hegde, Satish Mocherla, Joslyn K. Strebe, Xilong Li, Noelle S. Williams, Chao Xing, Mahmoud S. Ahmed, Ping Wang, Hesham A. Sadek, John W. Schoggins

**Author notes:** Author(s) of correspondence **Correspondence to: Mamta K. Jain**, MD, MPH, Professor of Internal Medicine, Division of Infection Diseases and Geographic Medicine, UT Southwestern Medical Center, **Hesham A. Sadek, MD, PhD**, Professor of Internal Medicine/Cardiology, Molecular Biology, and Biophysics, UT Southwestern Medical Center, John Schoggins, PhD, Associate Professor of Microbiology, UT Southwestern Medical Center.

## Abstract

**Background:** An in-silico screen was performed to identify FDA approved drugs that inhibit SARS-CoV-2 main protease (M^pro^), followed by in vitro viral replication assays, and in vivo pharmacokinetic studies in mice. These studies identified atovaquone as a promising candidate for inhibiting viral replication.

**Methods:** A 2-center, randomized, double-blind, placebo-controlled trial was performed among patients hospitalized with COVID-19 infection. Enrolled patients were randomized 2:1 to atovaquone 1500 mg BID versus matched placebo. Patients received standard of care treatment including remdesivir, dexamethasone, or convalescent plasma as deemed necessary by the treating team. Saliva was collected at baseline and twice per day for up to 10 days for RNA extraction for SARS-CoV-2 viral load measurement by quantitative reverse-transcriptase PCR. The primary outcome was the between group difference in log-transformed viral load (copies/mL) using a generalized linear mixed-effect models of repeated measures from all samples.

**Results:** Of the 61 patients enrolled; 41 received atovaquone and 19 received placebo. Overall, the population was predominately male (63%) and Hispanic (70%), with a mean age of 51 years, enrolled a mean of 5 days from symptom onset. The log_10_ viral load was 5.25 copies/mL vs. 4.79 copies/mL at baseline in the atovaquone vs. placebo group. Change in viral load did not differ over time between the atovaquone plus standard of care arm versus the placebo plus standard of care arm. Pharmacokinetic (PK) studies of atovaquone plasma concentration demonstrated a wide variation in atovaquone levels, with an inverse correlation between BMI and atovaquone levels, (Rho -0.45, p=0.02). In post hoc analysis, an inverse correlation was observed between atovaquone levels and viral load (Rho -0.54, p= 0.005).

**Conclusion:** In this prospective, randomized, placebo-controlled trial, atovaquone did not demonstrate evidence of enhanced SARS-CoV-2 viral clearance compared with placebo. However, based on the observed inverse correlation between atovaquone levels and viral load, additional PK-guided studies may be warranted to examine the antiviral effect of atovaquone in COVID-19 patients.

*clincialtrials*.*gov (NCT04456153)*.

## INTRODUCTION

SARS-CoV-2 was identified in late December 2019 as the causative agent of a severe acute respiratory syndrome named COVID-19[1-3]. Targeting the disease in the initial phase with an effective oral agent that can be used in the outpatient setting could mitigate the progression to severe disease and decrease need for hospitalization and mortality. As such, a large number of clinical trials have focused on testing a wide range of antiviral drugs against SARS-CoV-2. A clinicaltrials.gov search at the time this manuscript was written (May 2021), there were over 540 ongoing or completed clinical trials testing potential antivirals agents against SARS-CoV-2. These antivirals include new and repurposed drugs targeting viral proteins that are critical for viral replication such as the proteases (main protease (M^pro^) and papain-like protease) and RNA polymerase among others. For example, the first FDA approved antiviral drug against SARS-CoV-2 was remdesivir, which was originally developed for the Ebola virus, and has been successfully repurposed as a SARS-CoV-2 RNA polymerase inhibitor. Recently, Merck pharmaceuticals announced that Molnupiravir, an oral anti-viral agent, decreased the risk of hospitalization from COVID-19 by about 30% [4-6]. Also, Pfizer announced the clinical outcomes for their oral SARS-CoV-2 M^pro^ inhibitor, Paxlovid, that reduced risk of hospitalization or death by 89% (within three days of symptom onset) and 88% (within five days of symptom onset) compared to placebo[7, 8]. Both drugs were granted U.S. FDA Emergency Use Authorization. These new drugs however are unlikely to be widely available worldwide soon.

SARS-CoV-2 is an enveloped, single-stranded RNA betacoronavirus, with a genome size of 29,891 bases encoding for 29 proteins. The SARS-CoV-2 genome encodes several nonstructural proteins including main protease (M^pro^ or 3Cl^pro^), and papain-like protease (PLpro)[9]. Given the critical role that these proteins play in viral entry and replication, they have been the topic of intense bench and clinical studies. The first SARS-CoV-2 protein to be crystalized is the M^pro^ protein, which plays a critical role in generation of the viral proteome by cleaving viral polyproteins into individual proteins, resulting in generation of 12 non-structural proteins, including RNA-dependent RNA polymerase and helicase, which are required for viral replication. M^pro^ cleaves its target polypeptides after sequences that include the amino acid glutamine, and its substrate binding pocket is structurally unrelated to any human protease, and thus M^pro^ is a viable drug target for inhibition of SARS-CoV-2 replication [10-12].

A recent study performed an in-silico screen[13], followed by cell-based viral assays with authentic SARS-CoV-2 and identified several FDA approved drugs with antiviral activity. One drug, atovaquone, had an IC50 against SARS-CoV-2 that falls within its therapeutic range, although the antiviral effect of atovaquone does not appear to be primarily mediated by its M^pro^ inhibitory activity[13]. Based on the virocidal, pharmacokinetic and side effect profiles, as well as global drug availability, we chose atovaquone as a candidate for clinical testing. Here we report results of a 2-center, prospective, randomized, placebo-controlled clinical trial examining the antiviral effect of atovaquone in hospitalized COVID-19 patients.

## METHODS

### Design

This is a randomized, double-blind, placebo-controlled trial of atovaquone therapy in adult participants hospitalized with COVID-19. Enrollment into the trial began in July 22, 2020 and was completed on December 29, 2020 with 1 month follow-up completed January 26, 2021. There were two clinical trial sites located in Dallas, Texas Eligible participants were randomized in 2:1 fashion to atovaquone or matching placebo. The treatment group received atovaquone 1500mg BID PO for 10 days or matching placebo bid for up to 10 days, during hospitalization and after discharge. Participants could receive all available standard of care therapy under Emergency Use Authorization including remdesivir, dexamethasone and convalescent plasma as prescribed by the treating team. Atovaquone or placebo was administered orally or by nasogastric tube and was given with a meal or snack when possible. The trial protocol was approved by UT Southwestern Institutional Review Board and was overseen by an independent data safety and monitoring board, and all patients provided written informed consent. The trial was funded by a grant from UT Southwestern.

#### Eligibility

Patients were eligible if they had a positive polymerase chain reaction test for SARS-CoV-2 within 72 hours of hospitalization, ≥18 years of age, able to provide informed consent, and anticipated hospitalization for ≥48 hours. Patients were excluded if they met any of the following criteria: enrolled in another COVID-19 antiviral therapy, breastfeeding women, known hypersensitivity to atovaquone, treatment with rifampin, patients with AIDS who required treatment for *Pneumocystis jirovecii* or *Toxoplasma gondii*, not expected to survive for 72 hours, >14 days from symptom onset.

#### Randomized interventions

Atovaquone/placebo: Atovaquone and matching placebo were supplied by Pharmacy Solutions (Arlington, Texas).

#### Randomization

Randomization blocks of 12 were given separately to each site pharmacist and after a patient signed informed consent and eligibility was verified, a randomization code was given for each participant.

### Procedures

After randomization, 2 ml of saliva was collected from each participant and mixed with 2 ml of the preservative DNA/RNA Shield (Zymo Research) prior to administration of trial drug, and then repeated every evening and morning while the participant was in the hospital. Saliva, instead of nasopharyngeal swab, was collected because it provides a reliable viral load measurement,[14] and minimizes patient discomfort. Plasma and serum were collected at baseline (Day 1) and Day 3 and 5 of follow-up if still hospitalized. A telephone follow-up occurred at 2 and 4 weeks after randomization if the patient was discharged. All investigators remained blinded to study assignment until completion of follow-up and database lock. The lead investigators were involved in the design, analysis, and writing of the manuscript. Other investigators contributed in collection of data. All investigators reviewed the manuscript. The trial is registered on clincialtrials.gov (NCT04456153).

### RNA Isolation

Saliva was collected using the DNA/RNA Shield Saliva Collection Kit (Zymo Research) following the manufacturer’s protocol. 1-2 ml of saliva/Shield mix was incubated with DTT (Life Technologies) following the U.S. Department of Health and Human protocol (https://www.cdc.gov/coronavirus/2019-ncov/downloads/processing-sputum-specimens.pdf). The samples were then treated with Proteinase K (Zymo Research) following the manufacturer’s protocol. RNA was extracted using the Direct-zol RNA Miniprep Kit (Zymo Research).

### SARS-CoV-2 positive control

SARS-CoV-2 N gene was amplified from a synthesized N gene fragment (IDT) with primers that introduced a T7 promoter sequence on the 3’ end (IDT). The PCR product was purified using Qiagen PCR Purificaton Kit (Qiagen). In vitro transcription was performed using T7 RiboMAX Express Large Scale RNA Production System following the manufacturer’s protocol (Promega). RNA was quantitated by spectrophotometry on a DS-11 FX instrument (Denovix) and by fluorometer assay using the DeNovix RNA Assay. In vitro transcribed RNA was used to generate a standard curve for qPCR from a 10-fold dilution series starting at 5 × 10^10^ copies of RNA.

### RT-qPCR

RT-qPCR was performed in a 20 μl reaction containing 5 or 10 μl RNA, 5 μl TaqMan Fast Virus 1-Step Master Mix, 600 nM each primer, and 150 nM probe. 10 μl RNA was used for the detection of SARS-CoV-2 and 5 μl RNA for detection of GAPDH. SARS-CoV-2 primers and probe were designed as recommended by the Center for Disease Control (https://www.cdc.gov/coronavirus/2019-ncov/lab/rt-pcr-panel-primer-probes.html). GAPDH primers and probe were designed as previously reported[10]. All oligonucleotides were synthesized by LGC Biosearch Technologies. RT was performed at 50°C for 5 minutes, followed by inactivation at 95°C for 2 minutes, and 40 cycles of PCR (95°C for 3 seconds, 60°C for 30 seconds) on an ABI 7500 Fast thermocycler or a QuantStudio 3 (Applied Biosystems).

### Analysis of RT-qPCR data

For each 96-well plate, a standard curve of N gene dilutions was run as described above. A simple linear regression model was used to fit the Ct values from the standard curve and subsequently interpolate RNA concentrations from saliva samples. Across all 96-well plates, the R^2^ value for goodness of fit was 0.97 or higher. The lower limit of detection of the assay was 50ng N gene control RNA. GAPDH Ct values were obtained for all samples. Samples that had undetectable GADPH levels, a total of 7, were removed from the analysis. The GAPDH Ct values of the remaining samples were analyzed using the ROUT method for outlier analysis with a 1% threshold. This resulted in additional 4 samples being removed from the data set. The final data set contained 614 samples. SARS-CoV-2 RNA copies/mL were interpolated from the standard curves, and log-transformed data were analyzed.

### Outcomes and Statistical Analysis

Any sample with no detectable GADPH housekeeping and any with GADPH Ct >34 was prospectively omitted. Values below detection limit were assigned a value ½ between lowest detection limit and zero. All analyses were intention-to-treat.

The primary outcome was log transformed viral load (copies/mL) using generalized linear mixed-effect models of repeated measures (GLMM) using data from all samples and timepoints. Random intercepts with an unstructured covariance structure was used in the models. Time point differences were statistically assessed through contrast tests with the appropriate combination of the fixed effects of treatment group, time, and treatment group by time interactions. No adjustment was made for multiple comparisons. Statistical significance was set using alpha=0.05, and all analyses were performed using SAS version 9.4.

Secondary outcomes were (1) viral load (log copies/mL) at 2 days, 4 days, and 7 days after randomization. (2) Area under the curve (AUC) of viral load (log copies/mL) through day 3 and 7 using the trapezoidal rule. (3) Between group differences in viral load (log copies/mL) using GLMM stratified by morning and evening samples, use of remdesivir, median split of baseline values (high vs. low viral load), median split time from onset of symptoms (<5 days vs. ≥5 days, median split of body mass index (BMI), diabetes status, sex and age. (4) Time to 2 log unit decrease in viral load using Kaplan-Meier estimates.

Subgroup analyses evaluated the primary endpoint stratified by morning and evening samples, use of remdesivir, sex, diabetes status and median baseline viral load (high vs. low viral load), time from onset of symptoms (<5 days vs. ≥5 days), body mass index (BMI), and age. Our exploratory clinical outcome was to examine ≥2 point change in ordinal scale (where higher scores are associated with clinical improvement) at Day 5 by chi-square analysis as described previously[15].

### Pharmacokinetic Analysis

Blood was drawn from participants prior to drug being given on Day 1 (which is baseline). Patients received drug at approximately 5 pm on day 1 then twice per day thereafter (approximately 9 am and 5 pm). Blood was drawn on Day 3 in the morning and Day 5 in the morning (if patient was still in hospital) between 5 and 8 am.

Analysis of patient specimens utilized standard blood-borne pathogen precautions. Day 1 pre-dose samples were pooled and evaluated as blanks. Analysis was blinded to patient group, so samples from patients receiving placebo were analyzed in the same fashion as samples from patients receiving drug. Day 3 and 5 samples were diluted 1:25 or 1:50 in a total volume of 50 μL of commercial human plasma (BioIVT HMPLEDTA2, Lot BRH465874). To all samples, 10μL of internal standard (atovaquone-d4) diluted in 30 mM NH_4_ Acetate was added and samples vortexed. 400 μL of ethyl acetate was added to each sample. Tubes were vortexed for 30 sec, incubated at room temperature (RT) for 5 minutes and spun for 5 minutes at 16,100 x g. Supernatant was transferred to a second tube. To the first tube, 400 μL of ethyl acetate was added. Tubes were vortexed for 15 sec, incubated at RT for 5 minutes and spun for 5 minutes at 16,100 x g. Supernatant was removed and added to tube containing supernatant previously collected. Samples were dried down under vacuum and then resuspended in 100 μL of 20:80 dH_2_O:ACN, 5mM NH_4_ Acetate. Samples were vortexed for 15 seconds, sonicated for 3 minutes and spun for 5 minutes at 16,100 x g. Supernatant was transferred to an HPLC vial and analyzed on a Sciex 4000QTRAP coupled to a Shimadzu Prominence LC using a fit-for-purpose method. Atovaquone was detected in negative multiple reaction monitoring (MRM) mode using the following transitions: 365.096 to 337.0 (quantitation ion), 365.096 to 170.8 (qualifier ion). Atovaquone-d4 was detected using the 369.1 to 341.0 transition. An isocratic flow of 0.2 mL/min 2 mM NH_4_ acetate with 0.8 mL/min of acetonitrile on an Agilent C18 XDB column (5-micron, 50 × 4.6 mm) was used for chromatography. Atovaquone and Atovaquone-d4 showed a retention time of 1.73 min using this method. Concentrations were determined by comparison to a 9-point standard curve prepared by spiking blank human plasma with atovaquone standards made in DMSO. Standards and quality control (QC) samples were run twice with 14/18 standards and 7/8 QC’s showing back-calculated values with 15% of nominal. The limit of detection (LOD) was defined to be three-fold above the signal observed in blank plasma and the limit of quantitation (LOQ) was defined as the lowest point on the standard curve above the limit of detection and within 20% of nominal. The LOQ was 5 ng/mL. Recovery of analyte at low, medium and high concentrations was >93%. Final atovaquone levels were calculated as μg/ml at days 3 and day 5 following initiation of atovaquone therapy.

## RESULTS

Of the 61 patients who signed consents, 60 underwent 2:1 randomization; 41 were assigned to atovaquone group and 19 to the placebo group. Overall the population was predominately male and Hispanic with a mean age of 51 years. The two groups were balanced (Table 1) with regard to age, sex, race, co-morbidities, days from onset of symptoms, baseline oxygen requirements, and receipt of COVID-19 specific standard of care treatment. A higher proportion of participants with diabetes were in the atovaquone arm.

**TABLE 1:** Demographics and Clinical Characteristics at Baseline

|  | Overall<br>N(%) | Atovaquone<br>(n=41)<br>N (%) | Placebo<br>(n=19)<br>N(%) | P value |
| --- | --- | --- | --- | --- |
| **Gender** |  |  |  |  |
| Male | 38 (63) | 26 (63) | 12 (63) | 1.0 |
| **Race** |  |  |  |  |
| White | 46 (77) | 31 (76) | 15 (79) | 1.0 |
| Black | 8 (13) | 6 (15) | 2 (11) |  |
| **Ethnicity** |  |  |  |  |
| Hispanic | 42 (70) | 29 (71) | 13 (68) | 1.0 |
| Age, mean yeas (IQR) | 50.9 (41.9,<br>59.6) | 51.64 (42.5,<br>60.8) | 49.4 (41, 59.6) | 0.56 |
| BMI, mean | 32.78<br>(27,36.5) | 32.65(27.1,<br>35.9) | 33.07 (26.8,<br>37.1) | 0.86 |
| **Co-morbidities** |  |  |  |  |
| Hypertension | 38 (63) | 26 (63) | 12 (63) | 1.0 |
| Diabetes | 38 (63) | 30 (73) | 8 (42) | 0.04 |
| Obesity | 23 (38) | 15 (37) | 8 (42) | 0.78 |
| Chronic kidney disease | 20 (33) | 12 (29) | 8 (42) | 0.38 |
| Lung disease<sup>#</sup> | 12 (20) | 10 (24) | 2(11) | 0.31 |
| Heart disease<sup>^</sup> | 7(12) | 5 (12) | 2 (11) | 1.0 |
| Cancer | 6 (10) | 3 (7) | 3 (16) | 0.37 |
| transplant | 5 (8) | 2 (5) | 3 (16) | 0.31 |
| liver disease | 5 (8) | 4 (10) | 1 (5) | 1.0 |
| Vascular | 4 (7) | 2 (5) | 2 (11) | 0.23 |
| Other<sup>*</sup> | 3 (5) | 1 (2) | 2 (11) | 0.26 |
| **Other treatment** |  |  |  |  |
| On remdesivir |  | 27 (66) | 9 (47) | 0.26 |
| On dexamethasone |  | 30 (73) | 14 (74) | 1 |
| Plasma |  | 4 (10) | 1 (5) | 1 |
| **Other characteristics** |  |  |  |  |
| Days from symptom onset, mean days (IQR) | 5.15 (4,6) | 5.24 (4,7) | 4.95 (4,6) | 0.56 |
| **Oxygen status at baseline** |  |  |  | 0.57 |
| Room air | 17 (28.3) | 10 (24.4) | 7 (36.8) |  |
| Low flow oxygen | 40 (66.7) | 29 (70.7) | 11 (57.9) |  |
| High flow oxygen | 3 (5) | 2 (4.9) | 1 (5.3) |  |

<sup>#</sup> Chronic obstructive pulmonary disease, asthma, smoking
<sup>^</sup> Congestive heart failure, myocardial infarction, coronary artery disease, cardiomyopathy
<sup>\*</sup> Autoimmune, connective tissue, peptic ulcer, HIV\

### Primary Outcome

The log_10_ viral load was 5.25 copies/mL vs. 4.79 copies/mL at baseline in the atovaquone vs. placebo group and decreased in both groups over time. No differences in viral load over time were seen between the atovaquone plus standard of care arm versus the standard of care arm **(Figure 1A and 1B**).

**Figure 1.**
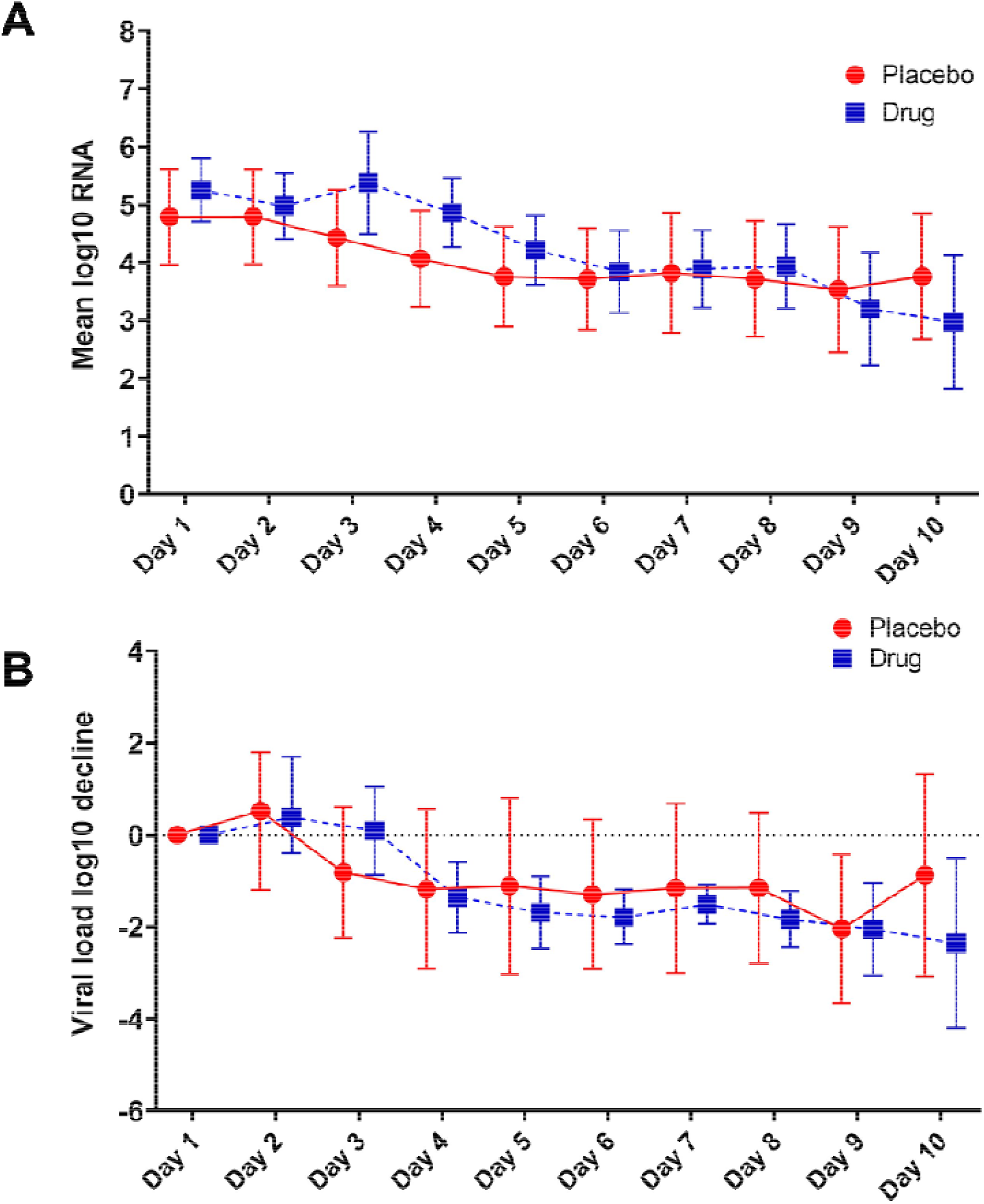
Primary Outcome. **A)** Mean log SARS-CoV-2 viral RNA in placebo compared to atovaquone over the 10-day trial period. **B)** Viral load decline in placebo and atovaquone groups. No statistically significant difference was detected between the two groups.

### Secondary Outcomes

Two days after intervention, the viral load was 5.37 copies/mL vs. 4.43 copies/mL in the atovaquone vs. placebo arm. Four days after intervention, the viral load was 4.22 copies/mL vs. 3.76 copies/mL and 7 days after the intervention, the viral load was 3.92 copies/mL vs. 3.71 copies/mL in atovaquone vs. placebo group. No differences were seen between the groups in any of the days. The AUC for viral load was 36.09 vs. 38.39, p=0.76 in the atovaquone compared with the placebo arm. There were no differences between groups in viral load over time in subgroup analyses stratified by sex, age, diabetes, time of sample collection, use of remdesivir, symptom onset of ≥5 days vs. <5 days, high versus low viral load. At Day 5, >2 point change in ordinal score occurred in 8 of 41 in atovaquone and 1of 19 in placebo, p=0.30. At Day 15, >2 point change in ordinal scale occurred in 25 of 40 in atovaquone and 9 of 17 in placebo, p=0.68.

### Atovaquone levels

Day 3 drug levels (7.668 μg/mL) were significantly lower than those on day 5 (11.590 μg/mL) (p =<0.01) **(Figure 2)**, suggesting that steady state plasma concentration was not reached by 3 days. Analysis of the correlation between BMI and drug levels revealed a statistically significant inverse correlation between BMI and atovaquone levels, (Spearman rho -0.45, p= 0.02) **(Figure 3)**. Interestingly, there was an inverse correlation between atovaquone levels and viral load (rho -0.54, p =0.005) **(Figure 4)**.

**Figure 2.**
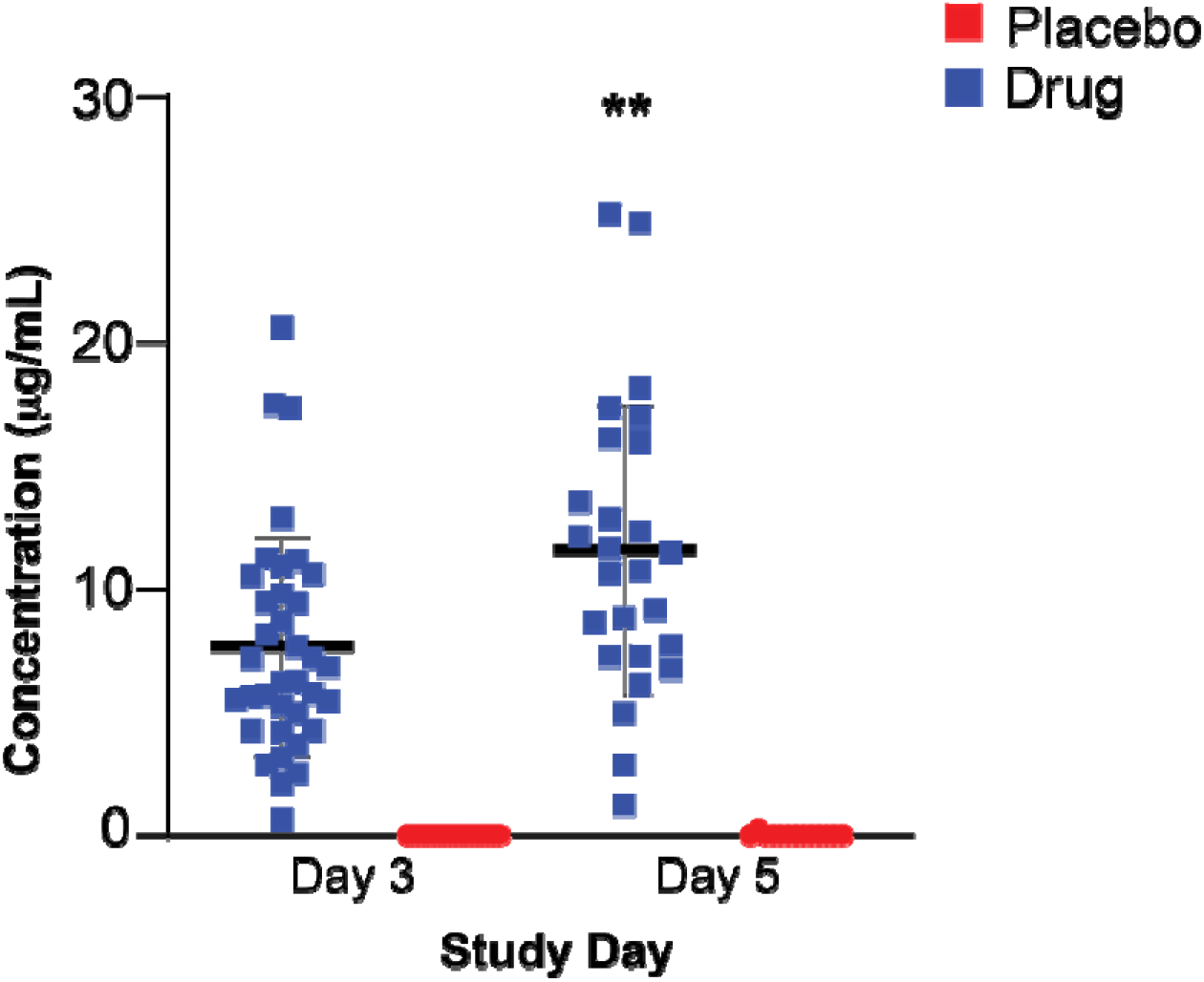
Atovaquone Plasma Concentrations. Atovaquone plasma concentration measured at day 3 and 5 following initiation of therapy. Day 3 drug levels were significantly lower than those of day 5. (** p<0.005).

**Figure 3.**
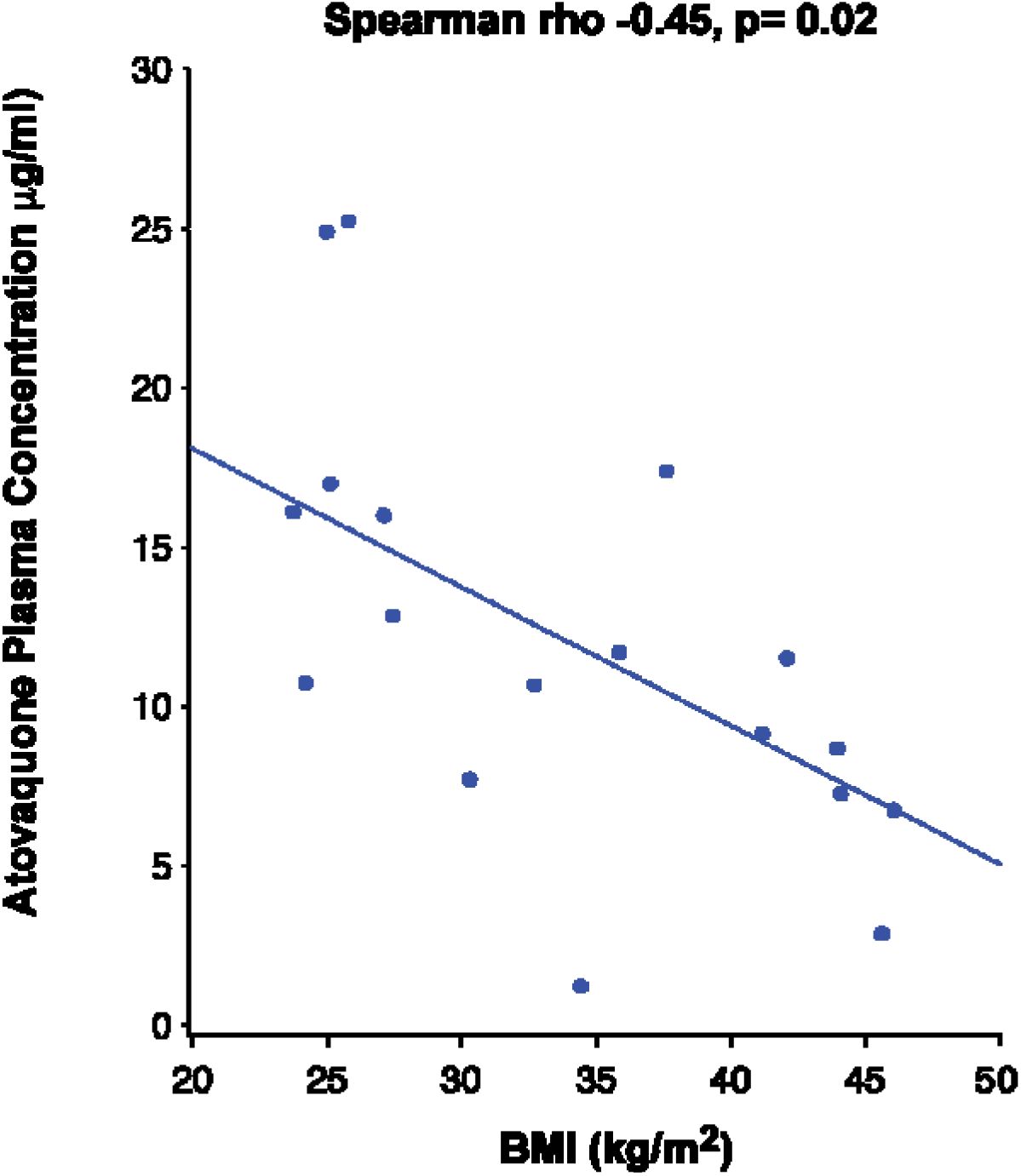
Correlation Between Atovaquone Plasma Concentration and BMI. Pharmacokinetic studies showed a negative correlation between atovaquone and BMI 5 days following initiation of atovaquone (rho -0.45, p-0.02).

**Figure 4.**
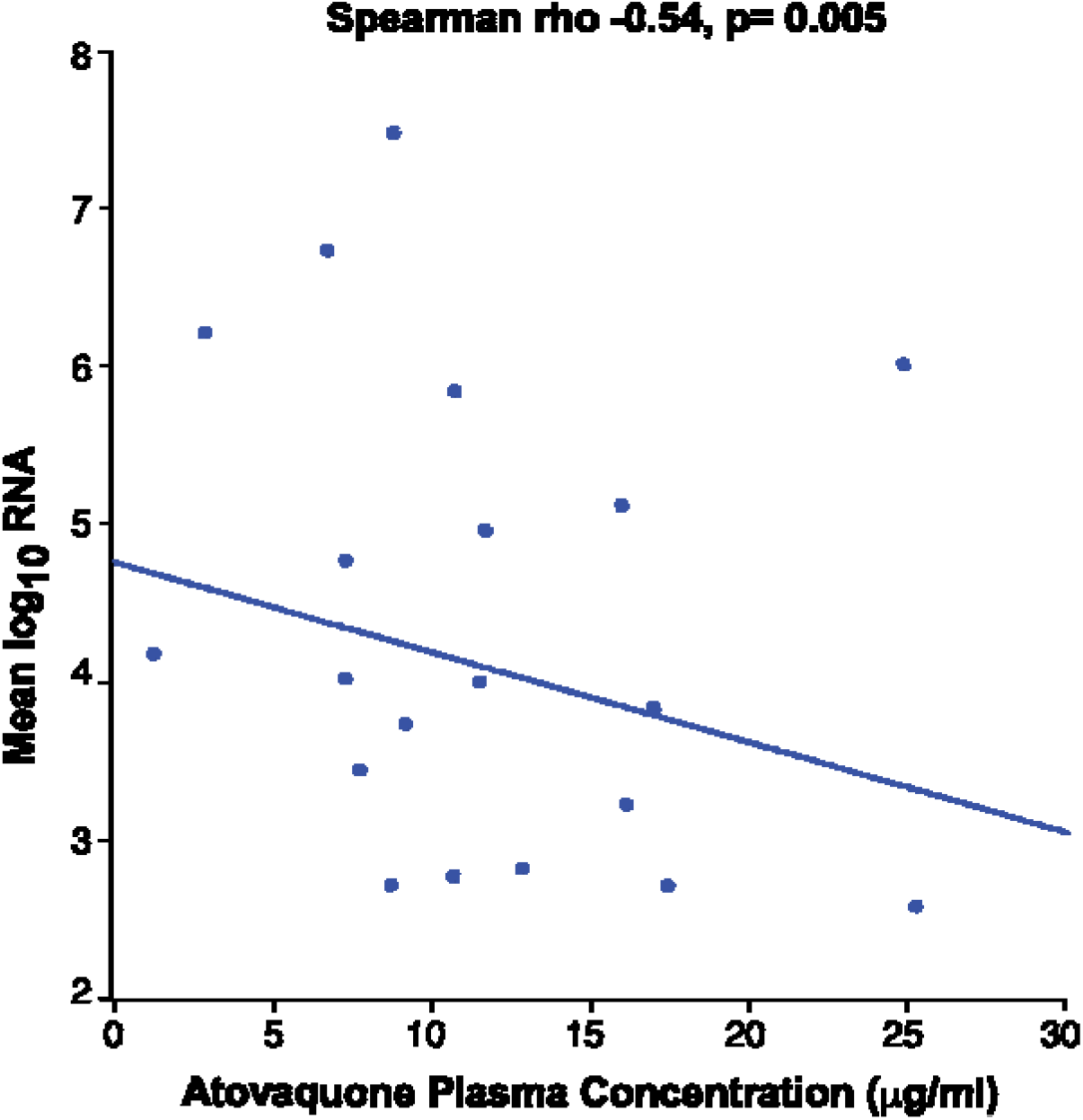
Correlation Between Atovaquone Plasma Concentration and Viral Load. Mean log SARS-CoV-2 viral RNA showed an inverse correlation with atovaquone plasma concentration (rho -0.54, p-0.005).

### Safety

There were 45 grade 3 or higher adverse events; 30 in the atovaquone arm and 15 in the placebo arm. There were two grade 3 adverse events thought to be related to study drug one in the atovaquone arm and one in the placebo arm. Non-serious adverse events thought to possibly related to study drug included hyponatremia, transaminitis, nausea/vomiting, and diarrhea. Overall, there was a total of 8 (13.3%) deaths in the trial, with 6 (14.6%) vs. 2 (10.5%), p=0.44 in the atovaquone vs. placebo group 28 days after intervention.

## DISCUSSION

In this prospective, randomized, placebo-controlled clinical trial of 60 patients, no effect on viral clearance was observed in patients treated with atovaquone compared with placebo. This trial was not powered to examine clinical efficacy. The results of present trial also demonstrated that atovaquone in patients hospitalized with COVID-19 was well tolerated. There was no significant difference in either severe adverse events or death in the atovaquone treated group.

The secondary outcomes and post-hoc analyses highlight several factors that may help explain why atovaquone did not show a significant effect on viral load compared with placebo as pre-specified in the primary outcome. First, the preclinical study that identified atovaquone did not test its antiviral effect *in vivo* in an animal model, and thus the *in vitro* antiviral effect in cell culture might not be directly translatable clinically. Second, atovaquone was administered in a hospitalized patient population, in which almost two-thirds received remdesivir as part of standard of care therapy, and thus it is possible that an antiviral effect of atovaquone was overshadowed by remdesivir. Importantly, although one-third of participants did not receive remdesivir, this study size was too small to allow exploration of the antiviral effects of atovaquone in patients not receiving remdesivir. It is also possible that measuring viral load in upper respiratory samples may not be a sensitive indicator for viral replication.

Another possibility for not detecting significant changes in viral kinetics in the overall population may be the inability to achieve inhibitory free drug concentration levels in some patients. Atovaquone has a long half-life and is highly plasma protein bound. The present study assessed the highest approved dose of 1500 mg twice daily. Based on PK/PD data from studies of *Pneumocystis jirovecii[16, 17]*, and PK studies in rodents[13], it was postulated that IC50-equivalent therapeutic plasma levels could be achieved. Our in vitro data indicated that the free atovaquone IC50 for SARS-CoV-2 antiviral activity in Vero cells is 1.2 nM based on a total IC50 of 1.5 μM and unbound fraction (f_u_) for atovaquone of 0.0008 in tissue culture media (N. Williams, unpublished). According to DrugBank, atovaquone is reported to be highly protein bound in plasma (>99.9%). Our preliminary analysis of atovaquone binding to human plasma supports this observation (f_u_ = 0.00003), suggesting that total drug levels of at least 40 μM (15 μg/mL) are needed. This, of course, assumes that the IC50 calculated in Vero cells is relevant for COVID-19 disease in vivo.

However, the PK data using samples collected from patients in the present trial indicate that IC50-equivalent drug levels were not achieved in most patients at 3 days after initiation of atovaquone, and only a subset of patients achieved adequate levels at day 5. Given that the trial was restricted to patients hospitalized with COVID-19, an antiviral effect during the early phase of the disease, when antivirals are most likely to have a therapeutic effect, was not tested in the current trial.

Importantly, the results highlight the potential role BMI may play with regard to atovaquone plasma concentrations. The present PK studies revealed two important findings: first, the inverse correlation between atovaquone plasma concentration and BMI is a strong indicator that patients with higher BMI may need higher dosing. Second, an inverse correlation was observed between atovaquone levels and viral load, which could suggest an antiviral effect of atovaquone on SARS-CoV-2 if adequate drug levels are achieved. However, the post-hoc nature of these findings preclude making reliable conclusions, and thus further PK-guided studies may be needed to determine the role of atovaquone in treatment of COVID-19 patients.

## Data Availability

All data produced in the present study are available upon reasonable request to the authors

## Acknowledgements

This trial was supported by funds from the Mary Kay Family Foundation and the Office of the President, UT Southwestern Medical Center. We are grateful for the following members of the Data Safety and Monitoring Board who reviewed the interim analysis: William M. Lee, MD; Kelly Chin, MD; Justin Grodin, MD; Joan Reish, PhD. We are grateful for Ezimamaka Ajufo, MD and Lorrie Burkhalter for developing data dictionary and RedCap instrument. The clinical team and pharmacy teams who assisted include: Tianna Petersen, MS; Laura Hansen, MS; Minerva Santos, Azadeh Mozaffari, Pharm D; Christine Cha, PharmD; Natalie Dellavalle; PharmD; Sonia Gonzales, PharmD. Pharmacokinetic analyses were conducted by the Preclinical Pharmacology Core at UT Southwestern Medical Center. HAS was funded by Center for Regenerative Science and Medicine at UT Southwestern Medical Center.

